# Most of *Staphylococcus aureus* and *Pseudomonas aeruginosa* co-infecting isolates coexist, a condition that may impact clinical outcomes in Cystic Fibrosis patients

**DOI:** 10.1101/2020.01.17.20017863

**Authors:** Paul Briaud, Sylvère Bastien, Laura Camus, Marie Boyadjian, Philippe Reix, Catherine Mainguy, François Vandenesch, Anne Doléans-Jordheim, Karen Moreau

## Abstract

*Staphylococcus aureus* (SA) is the major colonizer of the lung of cystic fibrosis (CF) patient during childhood and adolescence. As patient aged, the prevalence of SA decreases and *Pseudomonas aeruginosa* (PA) becomes the major pathogen infecting adult lungs. Nonetheless, SA remains significant and patients harbouring both SA and PA are frequently found in worldwide cohort. Impact of coinfection remains controversial. Furthermore, co-infecting isolates may compete or coexist. The aim of this study was to analyse if co-infection and coexistence of SA and PA could lead to worse clinical outcomes. The clinical and bacteriological data of 212 Lyon CF patients were collected retrospectively, and patients were ranked into three groups, SA only (n=112), PA only (n=48) or SA plus PA (n=52). In addition, SA and PA isolates from co-infecting patients were tested *in vitro* to define their interaction profile. Sixty five percent (n=34) of SA/PA pairs coexist. Using univariate and multivariate analysis, we confirm that SA patients have a clinical condition less severe than others, and PA induce a poor outcome independently of the presence of SA. FEV1 is lower in patients infected by competition strain pairs than in those infected by coexisting strain pairs compared to SA mono-infection. Coexistence between SA and PA may be an important step in the natural history of lung bacterial colonization within CF patients.

## Introduction

Cystic fibrosis (CF) is the most common genetic disease among Caucasian population that affects multiple organs and causes various complications associated with patient death such as cystic fibrosis liver disease (1) (CFLD) or cystic fibrosis related diabetes (2) (CFRD) (1, 2). However, the first cause of morbidity and mortality in CF remains the progressive decrease in pulmonary function leading to an obstructive syndrome (3). This decline is generally due to the continuous inflammation provoked by polymicrobial infections (3).

The main clinically significant bacteria are *Staphylococcus aureus* (SA) and *Pseudomonas aeruginosa* (PA). SA infection occurs early in children and affects up to 80% of patients aged 5-19 years [4]. In early childhood, SA infection is responsible for increasing inflammatory markers [5,6] and in adolescence, patients with high SA density in throat swabs present deterioration of lung function [7]. However, despite the implication of SA in worse clinical statue, PA, which becomes the dominant pathogen of the respiratory tract in adulthood [4] (up to 60% of patients > 18 years), remains traditionally the most feared pathogen due to its strong association with most severe clinical outcomes, such as a more robust inflammation, an increase in the number of exacerbations and a decrease in forced expiratory volume in one second (FEV1) [8-11].

Even if SA colonization decreases as patients aging, SA infection concerns more than 30% of CF adults (4). Among these cases, co-infection by PA and SA remains significant (5). However, it is difficult to determine whether this co-infection is a transitional stage between SA alone and PA alone or whether permanent co-infections with these two bacteria exist. Several *in vitro* studies deciphered the relationships between SA and PA in a context of pulmonary co-infection (6–10). According to recent studies, an evolution occurs regarding SA and PA clinical isolates interaction. PA early-colonizing isolates show a strong antagonism towards SA strains (7, 8) especially by producing anti-staphylococcal compounds, leading to a competition state (10). However, PA isolated from chronic lung infection lack this competitiveness and SA succeeds to coexist durably with PA (6, 9, 11). In this way, we previously demonstrated that in this context of coexistence without antagonism, the two bacteria may cooperate to persist more easily in lungs (Briaud *et al*., 2019).

Only few studies investigated the impact of SA-PA co-infection and clinical outcomes. In addition, none of them considered the interaction state (competition vs coexistence) between SA-PA. So far, available data have shown conflicting results on the link between SA-PA co-colonization and clinical outcomes. Ahlgren *et al*., did not find a significant clinical difference in adult patients co-colonized with SA plus PA compared to patient colonized solely by PA (12). Two studies highlighted a higher respiratory decline and rate of hospital admission for patients infected by PA alone in comparison with patients co-infected by SA-PA or by SA alone (5, 13). Finally, other studies reported that SA-PA co-infection is associated with a worse clinical outcome (14–17).

The aim of the study was to better characterize (i) co-infected patients with demographic data (age, BMI, gender) and (ii) the interaction state (competition or coexistence) between the two bacterial species. We also examined the consequence of SA-PA co-infection and SA-PA interaction state (competition vs coexistence) on pulmonary functions (FEV1%) and clinical outcomes (e.g. number of exacerbations, number of hospitalization). By computing demographic and clinical data with pulmonary infectious status, we gain more information about the impact of SA and PA infections and interactions in CF patient’s health.

## Materials and Methods

### Patients

The clinical and bacteriological data of CF patients supervised at the two CF Centers in Lyon, France, (CRCM: Centre de Ressources et de Compétences de la Mucoviscidose) were collected from February 2017 to August 2018.

The inclusion criterion was a stable microbiological status for SA and /or PA colonization, defined by the following criteria: i) at least three respiratory samples collected for each patient throughout the period considered (ii) at least two months between two successive samples (iii) all samples collected during the study for a patient had the same status with respect to the presence of SA and / or PA. The patients who did not match the criteria, or who were not co-colonized by SA or PA were excluded.

This study was submitted to the Ethics Committee of the Hospice Civil de Lyon (HCL) and registered under CNIL No 17-216. All patients were informed of the study and did not oppose to the use of their data.

### Clinical data gathering

Clinical data were extracted from computerized medical files (Easily®). Data collected were: gender, age at the time of the last sampling, CFTR genotype classified between severe and moderate genotype regardless of clinical severity (5, 18), pancreatic insufficiency defined by a faecal elastase < 200µg/g, CF-related diabetes (CFRD) or a carbohydrate intolerance, cirrhosis, an oral or enteral supplementation and/or an undernourishment (defined as a body mass index (BMI) (weight/height^2^) lower than 17kg/m2 in an adult and −2 standard deviations (SD) in a child according to the gender and age). The pulmonary function was defined by the FEV1 expressed as a percentage of the predicted value (%pred). We also gathered the number of hospitalizations, the length of hospital stays and the number of exacerbations during the 9 months preceding the last sample.

### Microbiology

The microbiological composition of each respiratory sample was determined by the Institute for Infectious Agent, HCL. The interaction state (coexistence or competition) of SA-PA pairs was defined by agar competition assay as previously described (Briaud *et al*., 2019). Briefly, SA and PA isolates recovered from co-infected patient’s expectorations were cultured in 10ml of BHI, at 37°C, 200 rpm. From overnight cultures, SA and PA suspensions were diluted to OD_600nm_ =0.5. Then, 100µL of SA suspension was spread uniformly onto trypticase soy agar (TSA) plates. After a drying time, 5µl of PA suspension were spotted at the centre of the plates. The plates were incubated at 37°C for 24 hours. The competitive phenotype was characterized by an inhibition halo of SA growth. In absence of inhibition halo, isolates were defined in coexistence.

Based on these microbiological analyses, the patients were categorized into four groups: (i) SA alone, (ii) PA alone, (iii) or SA-PA in competition, and (iv) SA-PA in coexistence.

### Statistical analysis

Two different analyses were performed using the same process: (i) SA vs PA vs SA+PA, and (ii) SA vs PA vs SA+PA in coexistence vs SA+PA in competition.

Factor Analysis of Mixed Data (FAMD) was used for initial data screen. Then, univariate analysis was done to determine significant differences between groups. For continuous variables (age, BMI, FEV1, number of hospitalizations, length of hospitalization and number of exacerbation), Kruskal-Wallis tests were used to identify whether a population was different from the others. Afterward, to individually test each pair of populations from significant Kruskal-Wallis tests, a Mann-Whitney Wilcoxon tests was used. Fisher’s exact tests were performed to compare categorical variables (CFTR genotype, gender, Denutrition, pancreatic insufficiency, CFRD, liver cirrhosis, enteral nutrition and food supply). For multiple comparisons, tests were corrected by a Bonferroni method and statistical significance was set with a q-value threshold at 0.05. Finally, a multinomial log-linear model (nnet package (19)) was used to determine significant associations between infectious status groups and variables retained by Akaike information criterion (AIC) (20). The AIC analysis was performed in order to identify the smallest and fittest set of variables describing our data. An adjusted odds-ratio with 95% confidence interval was reported for each final variable. All analyses were performed using R v3.5.3 (21).

## Results

### Clinical characteristics of patients

Of the 655 CF patients monitored in Lyon hospitals, we selected patients with at least three respiratory specimens with SA and / or PA during the study period. 276 patients were excluded because of the absence of the three required samples. Sixty-five patients had neither SA nor PA infection. Finally, 48 and 54 patients were excluded because of unstable SA-PA co-colonization during the period or the inability to assess their pulmonary function (age <4 years). Finally, 212 patients with CF were included in this study.

The cohort consisted of 124 adults and 88 children with a mean age of 21.70 years (range 4-69). The male-female ratio was homogeneous with approximately 51.42% (n=109) of men. About 80.66% (n=171) had a severe CFTR genotype and 41 patients had a moderate one. During this period, 39.62% (n=84) had an exacerbation and 25.94% (n=55) of the patients were hospitalized (Table S1).

The 212 patients were classified following their chronic bronchial colonization in 3 different groups: SA alone, PA alone and SA+PA. Co-infected patients were split into 2 subclasses (competition and coexistence) regarding the SA-PA interaction state determined by agar competition assay (Table 1). Among the 212 patients, 52.83% (n=112) had chronic bronchial colonization with SA, 22.64% (n=48) with PA and 24.53% (n=52) with SA+PA. Within the 52 co-infected patients, 65.38% (n=34) of SA and PA isolates presented a coexistence interaction and 34.62% (n=18) presented a competition phenotype. Patients solely colonized with PA were older (average of 32.02y) than co-infected patients (average of 23.38y) and themselves older than patients colonized by SA (average of 16.49y) (Table 1). This is consistent with the natural history of infections and international values (4).

**Table 1:** Clinical characteristics of CF patients according to their bacteriological status.

|  | SA alone | PA alone | SA and PA |  |  |  |
| --- | --- | --- | --- | --- | --- | --- |
|  | (%) | (%) | competition | coexistence | merge | p_value* |
| Number | 112/212 | 48/212 | 18/52 (34.61) | 34/52 (65.38) | 52/212 |  |
| Age (years) | 16.49 ± 8.77 | 32.02 ± | 23.67 ± 8.98 | 23.24 ± 10.52 | 23.38 ± | <0.0001<sup>a</sup> |
| ≥18 years | 45/112 | 42/48 | 14/18 | 23/34 | 37/52 |  |
| Sex, male | 63/112 | 23/48 | 10/18 | 13/34 | 23/52 | ns |
| Genotype |  |  |  |  |  |  |
| Moderate | 26/112 | 9/48 | 3/18 | 3/34 | 6/52 | ns |
| Severe | 86/112 | 39/48 | 15/18 | 31/34 | 46/52 |  |
| BMI (kg/m<sup>2</sup>) | 18.06 ± 2.68 | 20.74 ± 2.42 | 19.25 ± 2.56 | 19.71 ± 3.56 | 19.55 ± | <0.0001<sup>a</sup> |
| Undernourishment | 11/112 | 1/48 | 1/18 | 3/34 | 4/52 | ns |
| Food supply | 23/112 | 13/48 | 3/18 | 15/34 | 18/52 | ns |
| Enteral nutrition | 4/112 | 0/48 | 0/18 | 4/34 | 4/52 | ns |
| Pancreatic | 101/112 | 45/48 | 17/18 | 33/34 | 50/52 | ns |
| CF-related diabetes | 9/112 | 14/48 | 3/18 | 10/34 | 13/52 | 0.0007<sup>b</sup> |
| Cirrhosis | 6/112 | 2/48 | 0/18 | 1/34 | 1/52 | ns |
| Hospitalizations during |  |  |  |  |  |  |
| Number | 17/112 | 18/48 | 6/18 | 14/34 | 20/52 | 0.0003<sup>a</sup> |
| Length | 8.00 ± 6.10 | 17.39 ± | 7.33 ± 6.12 | 29.14 ± 27.39 | 22.60 ± | 0.0002<sup>a</sup> |
| Number of | 0.25 ± 0.69 | 1.44 ± 1.37 | 1.28 ± | 1.35 ± | 1.33 ± | <0.0001<sup>a</sup> |
| FEV1(% predicted) | 85.87 ± | 55.09 ± | 59.72 ± 18.73 | 64.59 ± 21.79 | 62.90 ± | <0.0001<sup>a</sup> |

BMI: Body Mass Index
FEV1: Forced Expiratory Volume in one second
\* P values from comparison between three groups: SA vs PA vs SA+PA (merge).
<sup>a</sup> P values from Kruskal-Wallis non-parametric test for comparison of the three groups (continuous variables)
<sup>b</sup> P values from Fisher's exact test for comparison of the three groups (categorical variables)

We proceeded to a FAMD analysis to decipher the similarity and heterogeneity between patient groups using both continuous and categorical variables. The two first retained axes accounted for 39.47% of the total variance of the data (Fig. S1). Four variables contributed to the first axe: number of exacerbations, number and days of hospitalization and FEV1. Age, BMI and not having pancreatic insufficiency contributed to the second axe. The repartition of the 212 patients based on the two first dimensions overlaped between the 4 groups (Fig. S1). Patients infected by SA were the most represented group and appeared to have clinical variable values more homogeneous than other patients. On the contrary, patients co-infected in a coexistence state seemed to have values with a greater heterogeneity (Fig. S1).

### Impact of the co-infection on CF patient clinical outcome

In order to define whether co-infection was associated with poorer clinical outcome, we compared three groups: patients infected with SA, patients infected with PA and patients co-infected with SA-PA. Continuous variable analysis differentiated the 3 patient groups (Table 2). SA patients seemed to be younger than co-infected patients, themselves younger than PA patients (p<0.0001). SA group stood out from others by lower BMI values (PA vs SA: p<0.0001, SA+PA vs SA: p=0.0074), a smaller number of hospitalizations (PA vs SA: p=0.0017, SA+PA vs SA: p=0.0013), a reduced length of hospitalizations (PA vs SA: p=0.0011, SA+PA vs SA: p=0.0010), a lower number of exacerbations (p<0.0001) and an higher FEV1 (p<0.0001). Co-infected and PA groups were not statistically different for number of hospitalizations, length of hospitalizations, number of exacerbations and FEV1. Only age (p=0,0012) and BMI (p=0,029) values were significantly different. For the analysis of categorical variables, CFRD was only statistically significant between PA versus SA mono-infected group (p=0.0031) and co-infected patients versus SA (p=0.0169) (Table S2).

**Table 2:** P-values for continuous variables comparisons between SA mono-infected (SA), PA mono-infected (PA) and co-infected groups (SA+PA). Mann-Whitney Wilcoxon non-parametric test corrected by a Bonferroni method was used.

|  | PA vs SA | SA+ PA vs<br>SA | SA+PA vs<br>PA |
| --- | --- | --- | --- |
| Age | <0.0001 | <0.0001 | 0.0012 |
| BMI | <0.0001 | 0.0074 | 0.029 |
| Number of hospitalizations | 0.0017 | 0.0013 | ns |
| Length of hospitalization | 0.0011 | 0.0010 | ns |
| Number of exacerbations | <0.0001 | <0.0001 | ns |
| FEV1 | <0.0001 | <0.0001 | ns |

To further investigate these results and to consider potential association between variables, multinomial analysis with infection type as outcome was performed using age, BMI, FEV1, number of exacerbations, number of hospitalizations, length of hospitalizations and CFRD (Table 3). Analyse through AIC criteria removed the three last characteristics suggesting that number and length of hospitalizations and CFRD could not be associated with infection type. The final model demonstrated that patients infected with PA were more likely to have a high BMI [OR (95% CI): 1.2662 (1.0313, 1.5545)], a higher number of exacerbations; [OR (95% CI): 2.0282 (1.2559, 3.2755)] and lower FEV1 [OR (95% CI): 0.9612 (0.9355, 0.9877)] relative to patients infected with SA (Table 3). The results were similar when the co-infected group was compared to SA only [OR (95% CI)]: BMI: 1.2022 (1.0015, 1.4432); exacerbations: 2.1660 (1.3514, 3.4078); FEV1: 0.9701 (0.9478, 0.9930)]. Comparing SA-PA group versus PA group, only age criteria was significant as co-infected patients were more likely to be younger than PA-mono-infected patients [OR (95% CI): 0.9477 (0.9017, 0.9961)].

**Table 3:** Adjusted odds ratios of cystic fibrosis patients’ infection status. Multinomial regression analyses were performed to study associations between the type of infection (SA mono-infected (SA), PA mono-infected (PA) and co-infected groups (SA+PA)) and clinical outcomes. Adjusted odds ratios with 95% confidence intervals (CI) are mentioned.

|  | PA vs SA |  | SA+ PA vs SA |  | SA+PA vs PA |  |
| --- | --- | --- | --- | --- | --- | --- |
|  | OR (95 % CI) | p_value | OR (95 % CI) | p_value | OR (95 % CI) | p_value |
| Age | - | 0.1354 | - | 0.6767 | 0.9477<br>(0.9017, 0.9961) | 0.0345 |
| BMI | 1.2662<br>(1.0313, 1.5545) | 0.0242 | 1.2022<br>(1.0015, 1.4432) | 0.0481 | - | 0.5913 |
| Number of<br>exacerbations | 2.0282<br>(1.2559, 3.2755) | 0.0038 | 2.1460<br>(1.3514, 3.4078) | 0.0012 | - | 0.7298 |
| FEV1 | 0.9612<br>(0.9355, 0.9877) | 0.0043 | 0.9701<br>(0.9478, 0.9930) | 0.0108 | - | 0.4947 |

### Impact of the bacterial coexistence and competition within co-infected CF patients

The second objective of this study was to determine whether the interaction profile between SA and PA could affect patient clinical outcome. Thus, we compared the mono-infected groups (SA only and PA only) with two co-infected groups: SA plus PA in coexistence and SA plus PA in competition.

Using continuous variables, the comparison between coexistence and competition groups showed no significant difference (Fig. 1). No statistical differences were also found between PA mono-infected and either SA plus PA in coexistence or in competition. Compared to SA groups, both SA plus PA interaction groups seemed to infect older patients (Coex. vs SA.: p=0.0057; Comp. vs SA: p=0.0243) with a higher number of exacerbations (Coex. vs SA: p<0.0001; Comp. vs SA: p=0.0003) and lower values of FEV1 (Coex. vs SA: p<0.0001; Comp. vs SA: p=0.0001). However, it is important to notice that, for BMI, number of hospitalization and length of hospitalization criteria, only coexistence group is significantly different from SA groups (Fig. 1). So, coexistence group presented higher values of BMI (p=0.0449), higher number of hospitalizations (p=0.0043) and longer stay of hospitalizations (p=0.0026) compared to SA groups.

**Fig 1:**
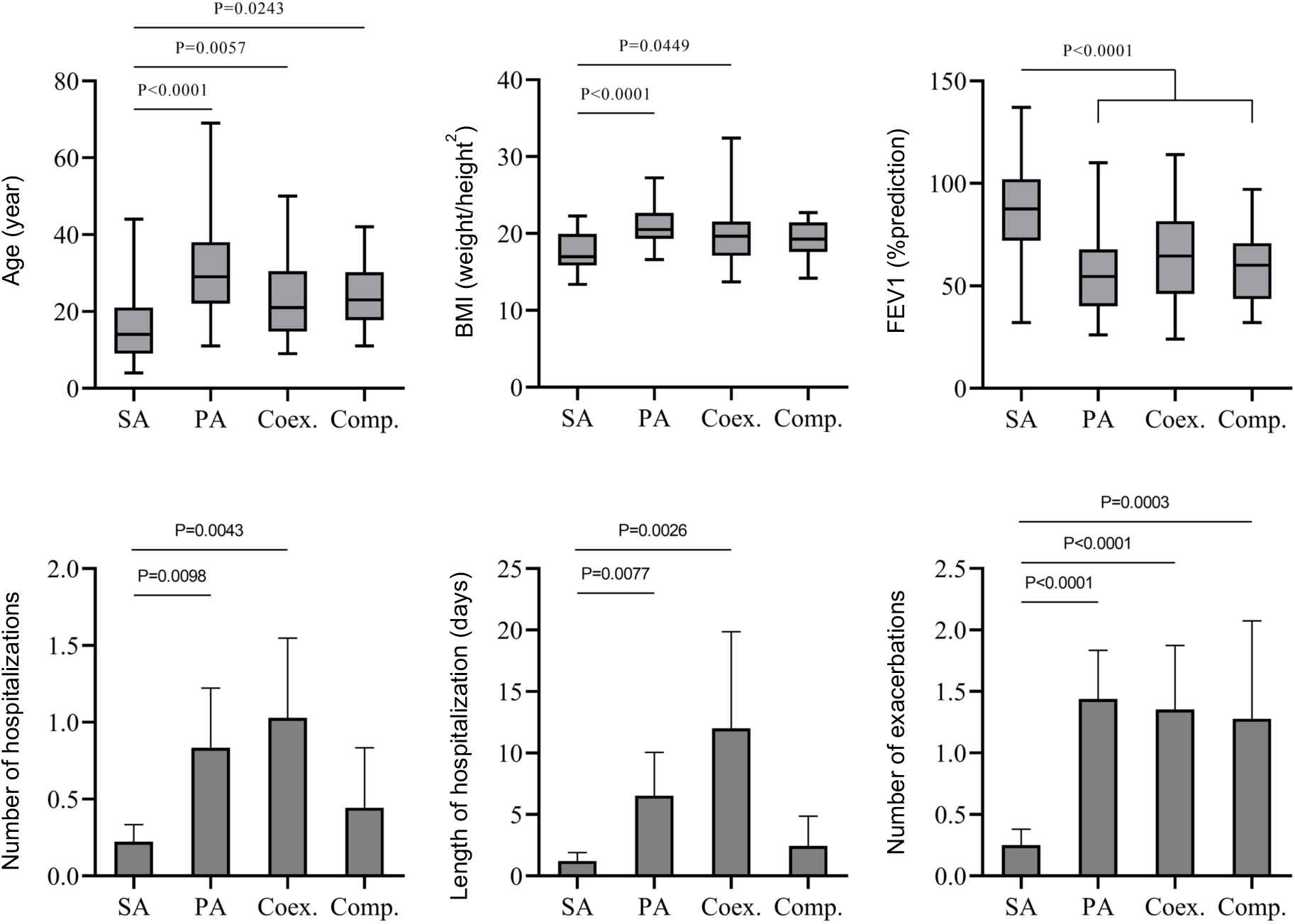
Comparison between SA mono-infected (SA), PA mono-infected (PA) and co-infected groups in competition (Comp) or in coexistence (Coex). Mann-Whitney Wilcoxon non-parametric test corrected by a Bonferroni method was used

Considering categorical variables, none characteristics were statically significant between the two types of interaction (Table S3). However, comparing SA group, there was higher number of CFRD patients in the coexistence group (p=0.0164); such criteria was not significant when SA group was compared to competition group.

Finally, multinomial analysis was performed using all significant characteristics from univariate tests. The final predictors retained after analyses of deviance through AIC criteria were BMI, supply of food, number of exacerbations and FEV1. A significant difference was found between the two types of interaction in co-infected patients. The coexistence state group was more likely to need food supply than the competition group [OR (95% CI): 0.2262 (0.0536, 0.9541)] (Table 4). As univariate results, multivariate analyses confirmed that patients co-infected by either a coexisting SA-PA pair, or a competition pair had higher odds of having more exacerbations than those infected only by SA [OR (95% CI): 2.2896 (1.3982, 3.7494): 1.9090 (1.0956, 3.3262) respectively]. Compared to SA group, the competition state group seemed to have lower values of FEV1 [OR (95% CI): 0.9589 (0.9300, 0.9887)] whereas the coexistence state group were more likely to have higher BMI [OR (95% CI): 1.2047 (1.0210, 1.4213)].

**Table 4:** Adjusted odds ratios of cystic fibrosis patients’ infection status. Multinomial regression analyses were performed to study associations between the type of infection (SA mono-infected (SA), PA mono-infected (PA) and co-infected groups in coexistence state (Coex.), or in competition state (Comp.)) and clinical outcomes. Adjusted odds ratios with 95% confidence intervals (CI) are mentioned.

|  | Coex vs SA |  | Comp vs SA |  | Comp Vs Coex |  |
| --- | --- | --- | --- | --- | --- | --- |
|  | OR (95 % CI) | p_value | OR (95 % CI) | p_value | OR (95 % CI) | p_value |
| BMI | 1.2047<br>(1.0210, 1.4213) | 0.0273 | - | 0.3199 | - | 0.4872 |
| Food supply | - | 0.0801 | - | 0.3609 | 0.2262<br>(0.0536, 0.9541) | 0.0430 |
| Number of exacerbations | 2.2896<br>(1.3982, 3.7494) | 0.0010 | 1.9090<br>(1.0956, 3.3262) | 0.0228 | - | 0.4472 |
| FEV1 | - | 0.0861 | 0.9589<br>(0.9300, 0.9887) | 0.0072 | - | 0.1885 |

### Diabetes outcome and infectious status are not associated

Additional multivariate analyses were performed concerning CFRD patients due to statistically significant difference between groups when performing univariate analyses (Table S2 and Table S3). First of all, a multinomial analysis was conducted with the entire variables (including the infection status) and taking CFRD as outcome. Through the analyses of deviance conducted by AIC criteria, the following variables were excluded from the multinomial analysis: sex, BMI, undernourishment, enteral nutrition, cirrhosis, length of hospitalisation, number of exacerbations, FEV1 and type of infection. Thus, all these variables were not associated to CFRD and CFRD patients were not more likely to be infected by neither SA nor PA nor coinfected by SA plus PA. Five characteristics were conserved in the final model where odds ratios and associated adjusted p_values were reported (Table 5). CF-related diabetes patients were more likely to be older [OR (95% CI): 1.1013 (1.0543, 1.1504)], to need more food supply [OR (95% CI): 3.0259 (1.2297, 7.4459)] and were more hospitalized [OR (95% CI): 2.1662 (1.5254, 3.0763)].

**Table 5:** Adjusted odds ratios of cystic fibrosis-related diabetes (CFRD) patients’ infection status. Multinomial regression analyses were performed to study associations between cystic fibrosis-related diabetes (CFRD) and other clinical outcomes including infection type. Adjusted odds ratios with 95% confidence intervals (CI) are mentioned.

|  | OR (95 % CI) | p_value |
| --- | --- | --- |
| Age | 1.1013<br>(1.0543, 1.1504) | <0.0001 |
| Genotype | - | 0.1022 |
| Food supply | 3.0259<br>(1.2297, 7.4459) | 0.0160 |
| Pancreatic<br>insufficiency | - | 0.8192 |
| Number of<br>hospitalizations | 2.1662<br>(1.5254, 3.0763) | <0.0001 |

## Discussion

Lungs of CF patients are colonized by multiple bacteria and several studies were conducted to decipher the impact of these colonisations on the clinical outcomes. We focused our study on the two major pathogens responsible for chronic colonization: *Staphylococcus aureus* and *Pseudomonas aeruginosa*. There is no gold standard to define chronic colonization especially for SA. For PA infection, Leeds criteria (22) are sometimes used and define ‘chronic’ when more than 50% of the preceding 12 months were PA culture positive. This definition requires frequent expectoration sampling. Furthermore, these criteria may lack sensitivity and are not always considered by clinicians (23). Alternatively, PA serology appears to be a sensitive criterion to classify PA infection, especially in non-expectorating children but is not always available (24). In order to clearly study the impact of SA and/or PA infection, we focused on stabilized infections. Therefore, we considered chronic colonization when at least 3 samples over a 9-month period had the same colonization pattern. These criteria are close to the definition of European Consensus Criteria for PA colonisation. However, this stringent method led to the exclusion of 33% of our cohort; 10% of patients were also excluded due to a change in colonization status during the period studied. Finally, in our study, 52.83% of the remaining patients had SA chronic colonization, 22.64% PA chronic colonization and 24.53% had chronic co-colonization with both bacteria. These data are consistent with previous studies (5, 25).

The first point of our study was to compare the impact of chronic colonization by SA alone, PA alone and SA plus PA on clinical outcome of CF patients. Indeed, being able to evaluate (or even predict) the risks that a patient incurs depending on the type of bacterium and/or on bacterial associations (in particular between PA and another pathogen) that colonize the lungs, remains one of the major challenges in the management of patients. Thus, identifying patients with harmful bacterial associations would be of clinical interest as they have increased risks of worse clinical outcome. These bacterial association could become a therapeutic priority and could be eradicated by targeting, for example, one of the pathogens. Conversely, we could speculate that co-infection between PA and another bacterium represents a milder step in the disease evolution, probably an intermediate stage between SA alone and PA alone.

In our study, we did not observe any differences regarding clinical status between patients co-infected by PA and SA and those infected by PA alone. This suggests that the SA plus PA combination does not lead to over- or under morbidity compared to the presence of PA alone in the lungs and that there is no addition of the effects of the two pathogens. As soon as PA colonise the CF lungs, whether SA is present or not, the clinical condition of the patients appears to deteriorate. This agrees with the study by Ahlgreen *et al*. [12] but disagree with others [5,14-17,22]. Indeed, studies on large size cohort with no distinction concerning age, suggested that PA mono-infected group was the most severe [5,22] unlike studies on smaller cohort focus on childhood concluded that co-infected group had the worst clinical outcomes [14,15,17]. In our study, we did not focus on specific age class due to the limited size of the cohort. The sizes of the cohorts and the age class studied could explain these dissimilarities and should be considered to analyse CF clinical outcome.

Our results showed that SA chronic colonization conducted to a higher FEV1 (85.87± 22.39), less exacerbations (0.25±0.69), a smaller number of hospitalizations (15.18% of SA patients) and shorter stay (8.00 ± 6.10) than SA plus PA co-infection or PA alone. This result confirms previous studies (12) showing that SA infection alone is a marker of milder disease comparing to PA, with less exacerbation rate and highest clinical score. Moreover, Hubert *et al*. (5) pointed out that co-infection by methicillin-susceptible SA (MSSA) and PA was more severe (lower FEV1 and higher IV antibiotic courses in one year) than mono-infection by MSSA. In the same way, Cios *et al*. (25) found that only MSSA group was milder comparing to co-infection group. Notably, association between SA alone and milder disease was not observed with MRSA. Indeed, MRSA was associated with higher hospitalization rate and re-hospitalization compared to co-infection. The SA methicillin resistant status seems to be, at the opposite, a severity marker of SA infection in CF patients. In our cohort, only 15 MRSA cases were reported. This low case number was not enough to perform analyses and conclude about impact of methicillin-resistance status on clinical outcomes. Further studies are required to figure out why MRSA - per se or as a surrogate marker of other genetic determinants-negatively impact the clinical outcome of CF patients. Moreover, other features of SA strains, such as the toxins production, could complement this analysis to more accurately describe the impact of SA in CF population.

CFRD is a severe complication of CF and association with bacterial infection could be used as a predictive marker of severity. CFRD seemed to be overrepresented in mono-infected PA (29.17%) and coinfected (25%) patients and under-represented in mono-infected SA patients (8.04%). CFRD represented 16.98% of our total patients and was slightly higher to CFRD rate in European population (26). Limoli *et al*. (16) showed that CFRD status was marginally associated, with an higher odds ratio, with co-infection by both SA and PA [OR (95% CI): 1.90 in comparison to SA or PA mono-infection(0,99, 3,6)]. We did not confirm this result. Indeed, our multivariate analysis demonstrate that CFDR was associated to the age of the patients [OR (95% CI): 1.1013 (1.0543, 1.1504)]. As co-infected and PA infected patients are older than SA patients, CFDR could be more represented in the two first groups. However, we cannot exclude that the difference between the two studies can be explained by a lower number of patients in our study.

The second objective of the present study was to evaluate the impact of the type of interaction between SA and PA (competition or coexistence) on clinical outcomes. Among the 52 patients co-colonized with SA and PA, 65.38% of strain pairs were in coexistent interaction and 34.61% in competition. These competitive and coexistence status were already described (6–8, 11, 27, 28). However, this is the first time that the type of interaction was evaluated in a large number of clinical isolates and we described that coexistence is the predominant interaction type in chronically co-colonized patients.

It has been accepted and described for a long time that PA could suppress SA growth by several mechanisms (10). These data led to the theory that PA colonization causes SA elimination into lungs patients. In fact, in our cohort, 24% of patients are co-colonized, which is consistent with other studies (5, 16). Moreover, SA-PA pairs from co-infected patients were mostly coexisting. Thus, our study shows that in 34 patients, chronical co-colonization may be due, at least in part, to the inability of PA to suppress SA. It has been well described that, in CF lungs, PA strains evolve and adapt from virulent early-infecting strains to less-virulent late-infecting strains (6, 8). Stresses induced by the pulmonary environment (oxidative and osmotic), antibiotic treatments, immune system and other bacteria species, constitutes selective pressure forces for adaptive mutations selection (8, 11, 17). Virulence changes could also reduce the antagonistic action of the PA isolates on SA (7, 8). However, of all the factors affecting PA’s evolution to the pulmonary environment, the role of SA on PA evolution remains to be explored.

Nonetheless, in the present study, we did not observe a different age between patients colonized with coexisting isolates (23.24 ± 10.52) and patients colonized with competitive pairs (23.67 ± 8.98). This suggests that there would be no link between adapted late-infecting strains and coexistence. In order to clarify this point, longitudinal studies will be necessary to evaluate the link between bacteria interaction status and isolates evolution.

How PA tolerance towards SA may contribute to worst clinical outcome is a point that has not been explored before. Except for food supply through multinomial analyses, no significant difference in clinical outcomes between patient with coexisting pairs and patients with competitive pairs was observed. Nevertheless, compared to SA group, patients infected by competition pair were more likely to have lower FEV1 than those infected by coexisting pair. These results may suggest that competition isolates may induce a more severe disease. Several quorum sensing dependent virulence factors are produced by competition PA strains such as rhamnolipids and phenazines (10), that could trigger an higher inflammatory response and a lower FEV1. On the contrary, coexisting PA strains lacking the production of virulence factors could less damage the lung environment. The clinical outcome is therefore mainly due to the genotype/phenotype of PA. However, as described previously, the PA adaptation is related to many environmental factors, including the surrounding bacteria. Thus, in the coexistence context, a current analysis of SA impact on PA transcriptome and growth phenotype suggests that SA could favour PA persistence (Camus et al., in preparation), and then influence the clinical evolution. A larger patient cohort would be necessary to evaluate the real impact of coexistence state on clinical outcomes. Coexistence between SA and PA may be an important step of lung bacterial colonization in CF patient, and preventing this phenomenon could participate in an adapted or even personalized management of CF patients.

### Ethical statement

All methods were carried out in accordance with relevant French guidelines and regulations. This study was submitted to the Ethics Committee of the Hospices Civils de Lyon (HCL) and registered under CNIL No 17-216.

## Data Availability

all data may be obtained on request to corresponding author

## Acknowledgments

“Vaincre la mucoviscidose” and “Gregory Lemarchal” associations supported this work. P Briaud was supported by French Ministry of Education and Research and L Camus was supported by Fondation pour la Recherche Médicale (grant number ECO20170637499). We thank Marie Verneret for technical support.

## Authors’ contributions

PB, ADJ and KM contributed to conception and design of the study. PR, CM and FV contributed to design of the study. PB and LC conducted the experiments. SB conducted and analyzed all statistics. MB and ADJ collected the data. All authors contributed to manuscript and approved the submitted version.

None of the authors have any conflict of interest to report.

## Supplementary data

**Fig. S1:**
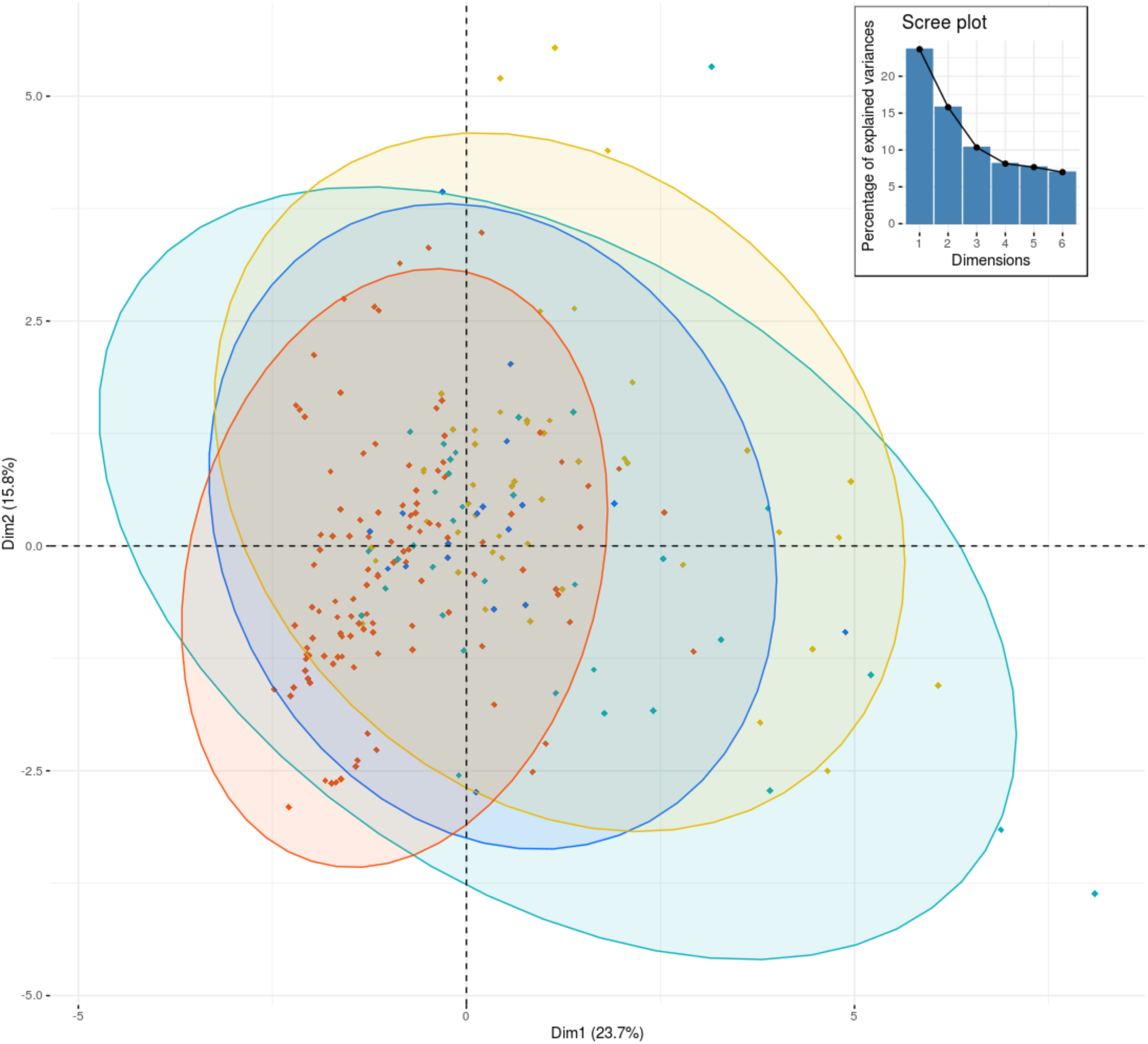
Individual factor map from FAMD based on 212 patients using the two first principal components. Patients are colored according to their infection type, SA mono-infected (red), PA mono-infected (yellow), SA+PAcoinfection in a competition state (blue), SA+PA coinfection in a coexistence state (cyan). Associated eigenvalue plot is shown at the top right.

**Table S1:** Patients characteristics at the time of the study.

| | Numbers of | Mean ± SD or |
| --- | --- | --- |
| Baseline characteristics | patients | Percentage |
| Age (years) | 212 | 21.70 ± 12.09 |
| $\geq 18$ years | 124/212 | 58.49% |
| Sex, male | 109/212 | 51.42% |
| Genotype |  |  |
| Moderate | 41/212 | 19.34% |
| Severe | 171/212 | 80.66% |
| BMI (kg/m<sup>2</sup>) | 212 | 19.03 ± 2.97 |
| Undernourishment | 16/212 | 7.55% |
| Food supply | 54/212 | 25.47% |
| Enteral nutrition | 8/212 | 3.77% |
| Pancreatic sufficiency | 16/212 | 7.55% |
| CF-related diabetes | 36/212 | 16.98% |
| Cirrhosis | 9/212 | 4.25% |
| Hospitalizations during |  |  |
| study period | 55/212 | 25.94% |
| Number | | 1.96 ± 1.17 |
| Length (days) | | 16.38 ± 18.28 |
| Exacerbations* | 84 | 0.78 ± 1.24 |
| FEV1 (% predicted) | 212 | 73.27 ± 25.12 |

BMI: body mass index
FEV1: forced expiratory volume in one second
\* Corresponding to total number of exacerbations

**Table S2:** P-values for categorical variables comparisons between *S. aureus* mono-infected (SA), *P. aeruginosa* mono-infected (PA) and co-infected groups (SA+PA). Fisher’s exact test corrected with a Bonferroni method was used.

|  | PA vs SA | SA+ PA vs SA | SA+PA vs PA |
| --- | --- | --- | --- |
| Sex | - | - | - |
| Genotype | - | - | - |
| Undernourishment | - | - | - |
| Food supply | - | - | - |
| Enteral nutrition | - | - | - |
| Pancreatic sufficiency | - | - | - |
| CF-related diabetes | 0.0031 | 0.0169 | ns |
| Liver cirrhosis | - | - | - |

**Table S3:** P-values for categorical variables comparisons between *S. aureus* mono-infected (SA), *P. aeruginosa* mono-infected (PA), co-infected in a coexistence (Coex) state and co-infected in a competition (Comp) state group.

|  | Coex vs<br>SA | Comp vs<br>SA | Coex<br>vs PA | Comp<br>vsPA | Comp.<br>vs<br>Coex |
| --- | --- | --- | --- | --- | --- |
| Sex | - | - | - | - | - |
| Genotype | - | - | - | - | - |
| Undernourishment | - | - | - | - | - |
| Food supply | ns | ns | ns | ns | ns |
| Enteral nutrition | - | - | - | - | - |
| Pancreatic<br>insufficiency | - | - | - | - | - |
| CF-related<br>diabetes | 0.0164 | ns | ns | ns | ns |
| Liver cirrhosis | - | - | - | - | - |

